# Pharmacokinetics, target attainment and outcomes of piperacillin/tazobactam in critically ill patients receiving continuous infusion with therapeutic drug monitoring: a retrospective analysis

**DOI:** 10.64898/2025.12.18.25342590

**Authors:** Ute Chiriac, Max Münchow, Anka C. Röhr, Otto R. Frey, Anna T. Frey, Daniel Frisch, Maxime Gaasch, Markus A. Weigand, Sebastian G. Wicha, Alexander Brinkmann

**Affiliations:** Hospital Pharmacy, Heidelberg University Hospital, Heidelberg, Germany; Department of Clinical Pharmacy, Institute of Pharmacy, University of Hamburg, Hamburg, Germany; Department of Clinical Pharmacy, General Hospital of Heidenheim, Heidenheim, Germany; Department of Anesthesiology, General Hospital of Heidenheim, Heidenheim, Germany; Department of Anesthesiology, Heidelberg University Hospital, Heidelberg, Germany

**Keywords:** piperacillin/tazobactam, continuous infusion, therapeutic drug monitoring, pharmacokinetics, critically ill patients

## Abstract

**Objectives:** To provide real-world evidence on piperacillin exposure and outcomes in critically ill patients following the implementation of pharmacokinetic (PK)/pharmacodynamic (PD)-guided dosing in routine care.

**Methods:** This retrospective observational study included critically ill adults who received continuous piperacillin/tazobactam infusion between 2011 and 2019. Empiric doses were individualized using dosing software based on renal function and subsequently adjusted according to therapeutic drug monitoring (TDM) results. Drug exposure was defined as subtherapeutic (<32 mg/L), therapeutic (32–64 mg/L), moderately high (64–96 mg/L), or supratherapeutic (>96 mg/L).

**Results:** A total of 1538 critically ill patients with severe infections and sepsis of varying severity were included, and 3,090 piperacillin serum concentrations were analysed. Median daily piperacillin dose was 8,000 mg, median steady-state concentration 55 mg/L, and median clearance 6.25 L/h. At the first measurement, individualized empiric dosing resulted in 45.7% of patients being within the therapeutic range; after TDM-guided adjustment, target attainment increased to 62.4%. Subtherapeutic and supratherapeutic concentrations were uncommon among all TDM samples collected during individualized dosing (<32 mg/L: 12.8%; < 16 mg/L: 0.8%; > 96 mg/L: 11%). ICU mortality was 21.1 % in patients within the therapeutic range, 30.3 % in those with moderately high concentrations, and 44.1 % in those with supratherapeutic concentrations (p = 0.05). Women were 1.8 times more likely to present supratherapeutic concentrations.

**Conclusions:** A multimodal approach combining individualized empiric dosing, TDM, and continuous infusion ensured target attainment while reducing drug consumption. These findings support the integration of individualized, PK/PD-guided dosing into routine care for critically ill patients and highlight the need for further studies addressing sex-related pharmacokinetic variability.

## Introduction

Despite increasing emphasis on precision medicine, antibiotic dosing in critically ill patients often still follows standardized regimens. This approach does not adequately account for the complex pathophysiological changes during sepsis and septic shock [1], leading to substantial variability in beta-lactam concentrations [2–4]. Observational studies demonstrated that standardized regimens achieve conservative pharmacokinetic (PK)/pharmacodynamic (PD) targets for beta-lactams, defined as maintaining concentrations above the minimum inhibitory concentration (100% *f*t>MIC), in only about 60 % of critically ill patients [2–4].

To address this challenge, different strategies have been suggested to tailor antibiotic dosing to individual patient characteristics and antibiotic-specific PK/PD properties. Prolonged (extended or continuous) infusion improves target attainment by maintaining concentrations above the MIC, thus reflecting their time dependent bactericidal activity. The BLING III trial, a large prospective multicentre RCT, demonstrated significantly higher clinical cure rates with continuous infusion [5]. When pooled with other studies in a recent meta-analysis, prolonged infusion was associated with improvements in both mortality and clinical cure, thereby highlighting continuous infusion as an increasingly accepted standard of care in critically ill patients [6–8].

Therapeutic drug monitoring (TDM) enables real-time adjustment of dosing to achieve adequate exposure. The TARGET trial demonstrated improved target attainment with TDM but no mortality benefit [9], whereas post-hoc machine learning analysis suggested possible improvements in clinical recovery [10]. In addition, dose individualization at treatment initiation, e.g., supported by dosing software, can help to achieve adequate exposure from the very beginning of therapy, even before TDM results are available [11]. A recent meta-analysis indicated that individualized dosing, using TDM and/or dosing software, was significantly associated with higher target attainment and lower rates of treatment failure and nephrotoxicity, while trends towards improved survival and clinical cure did not reach statistical significance [12]. Furthermore, an individualized approach might also have economic implications, as it could lead to a significant reduction in therapy costs for critically ill patients [13, 14].

In 2009, an individualized, PK/PD-guided approach was implemented for piperacillin/tazobactam therapy in the intensive care unit of the Department of Anaesthesiology, General Hospital of Heidenheim. This retrospective analysis evaluates piperacillin exposure and outcomes to provide real-world evidence and to support the integration of individualized dosing, TDM, and continuous infusion into routine practice.

## Methods

### Study Setting and Population

This retrospective observational study was conducted at a German academic teaching hospital. Ethical approval was waived by the Ethics Commission of the University of Ulm. Data from 2013 have been reported previously [15]; the present analysis includes follow-up data from 2011 and 2019. The study included critically ill adults with sepsis, severe sepsis, or septic shock who received empiric piperacillin/tazobactam by continuous infusion. Children or those treated with intermittent infusion were excluded.

### Therapeutic range and definition of exposure

The therapeutic range was defined as piperacillin concentrations between 32 and 64 mg/L, which corresponds to 2 to 4 times the MIC breakpoint for *Pseudomonas aeruginosa* and 4 to8 times the MIC breakpoint for *Enterobacterales* and anaerobic bacteria[*16*]. Concentrations between 64 and 96 mg/L were classified as moderately high, while concentrations above 96 mg/L were defined as supratherapeutic. Piperacillin clearance (CL*_PIP_*) was estimated from observed steady-state concentrations and the utilised infusion rate. Concentrations for alternative dosing regimens (16 g per day as recommended by the German Interdisciplinary Association for Intensive Care and Emergency Medicine (DIVI) [17], standard dosing according to the Summary of Product Characteristics (SmPC) [18]) were predicted accordingly. Renal function was assessed using the Cockcroft–Gault equation; adjustments for low serum creatinine and use of adjusted body weight followed standard practice. Details are described in the online supplement.

### Antibiotic Administration, TDM, and Data collection

Antibiotic treatment was initiated within the first three hours of diagnosis, in accordance with the Surviving Sepsis Campaign (SSC) guidelines [19]. Piperacillin/tazobactam was administered as a continuous infusion, beginning with a loading dose of 2000/250 mg. Empiric maintenance dose were calculated using the CADDy software (www.thecaddy.de) based on renal clearance and, if applicable, dialysis settings [1]. Dose adjustments were performed within the first 24 to 48 hours based on TDM results under supervision of trained clinical pharmacists. Piperacillin concentrations were determined using a validated high-performance liquid chromatography (HPLC) assay [20].

### Statistical Analysis

Statistical analyses were performed using the R (version 4.5.0, R Core Team) and RStudio (version 2025.05.0+496, Posit team) software. Continuous variables were summarized as mean ± standard deviation (SD) or median with interquartile range (IQR). Categorical variables were presented as counts and percentages. Group comparisons for continuous variables were performed using the Wilcoxon rank-sum test. For binary or categorical variables, Fisher’s exact test was applied. Spearman’s rank correlation was used to assess associations between continuous variables; a correlation coefficient (ρ) > 0.7 was considered strong. To evaluate the relationship between piperacillin plasma concentration category and ICU mortality, a binary logistic regression model was used with a logit link function. Odds ratios (ORs) with 95% confidence intervals (CIs) were reported to quantify the strength of association between initial exposure and therapeutic outcome (mortality). A second model adjusted for SAPS II score to account for baseline severity of illness. Initial piperacillin concentration category and its association with continuous covariates (e.g., renal clearance, dose) were further explored using generalized linear models (GLMs). Model type and link function were selected based on the distribution of the outcome variable. To control type I error inflation due to multiple comparisons, p-values were adjusted using the Holm–Bonferroni method. A two-sided p-value ≤ 0.05 was considered statistically significant. No imputation was performed for missing data; all analyses were conducted using complete cases.

## Results

### Patient characteristics

In total, 1538 critically ill patients were included, representing a typical patient population in a German intensive care unit. All patients presented with severe infections. Pneumonia (65.8%) and abdominal infections including peritonitis (17.9%) were the most common infection foci, while urinary tract infections (9.4%) and other sites (6.8%) were less frequent. The duration of piperacillin/tazobactam therapy ranged from 1 to 20 days (median 7 days). Detailed demographic and clinical characteristics are summarized in Table 1.

**Table 1:**
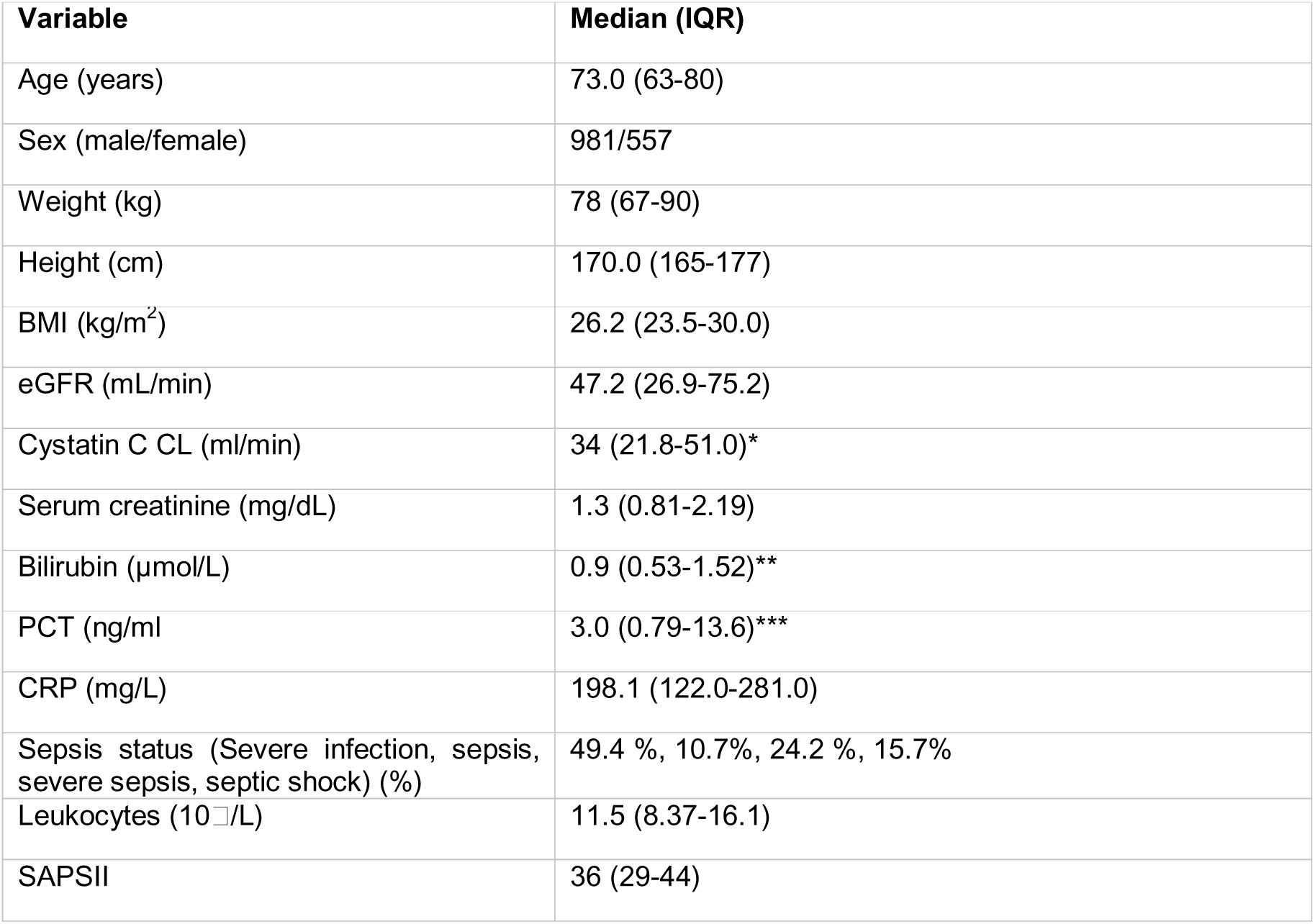
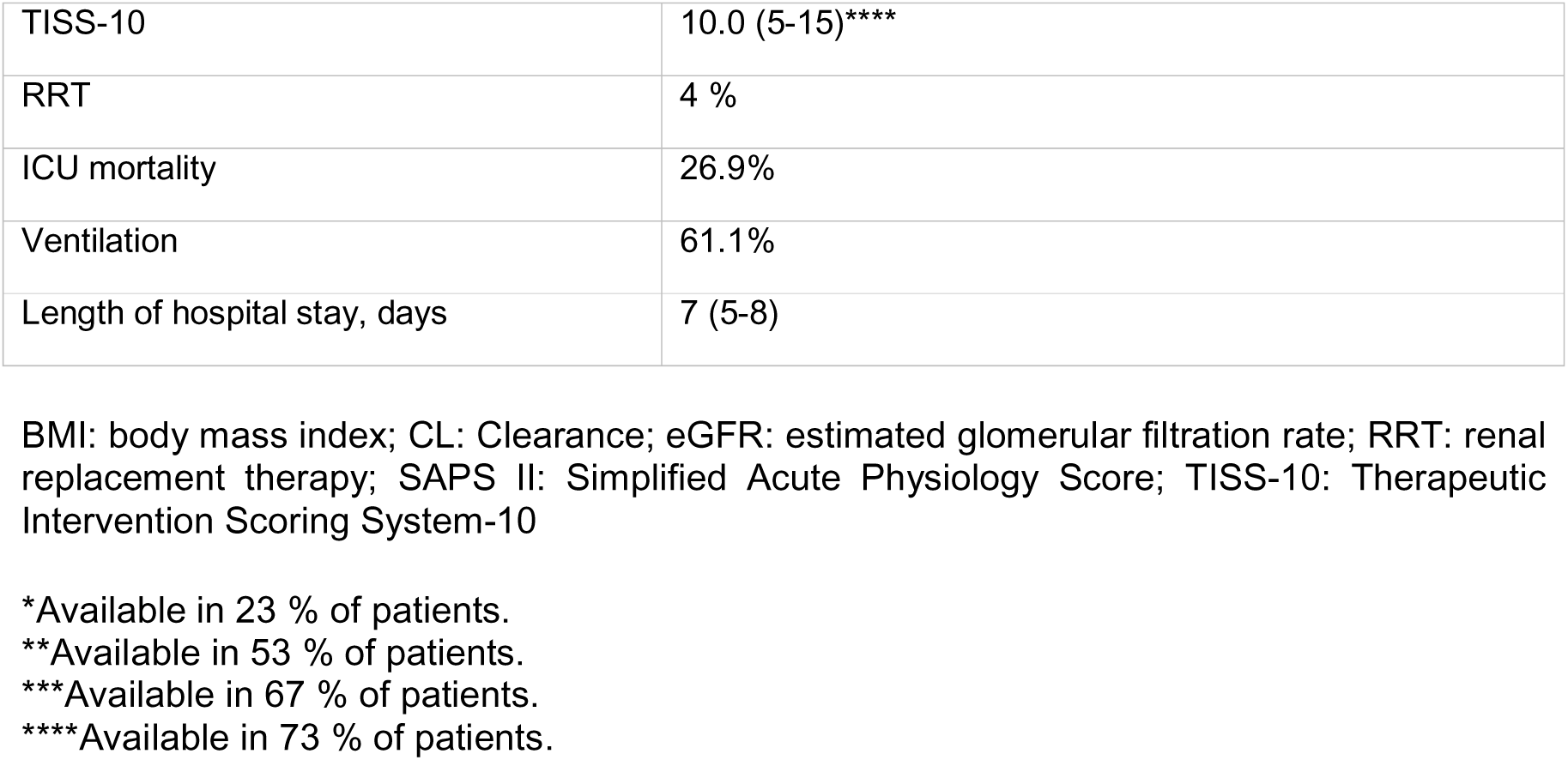
Baseline demographic and clinical characteristics.

### Piperacillin exposure

A total of 3,089 piperacillin concentrations were analysed. The median concentration was 55 mg/L (range 11-290 mg/L) (see web-only Supplementary Figure S1). Piperacillin clearance showed a wide interindividual variability, with a median of 6.25 L/h (range 0.65–55.09 L/h) (Figure 1). Similarly, individual maintenance doses varied from 1.4 g/24 h to 24.2 g/24 h, with a median of 8.0 g/24 h (Figure 2). Observed target attainment and predicted attainment for the alternative dosing strategies are presented in Table 2 and in web-only Supplementary Figure S2. Subtherapeutic concentration measurements were rare among all TDM samples, and only a small subset of these were below 16 mg/L (26/3089, 0.8%).

**Figure 1:**
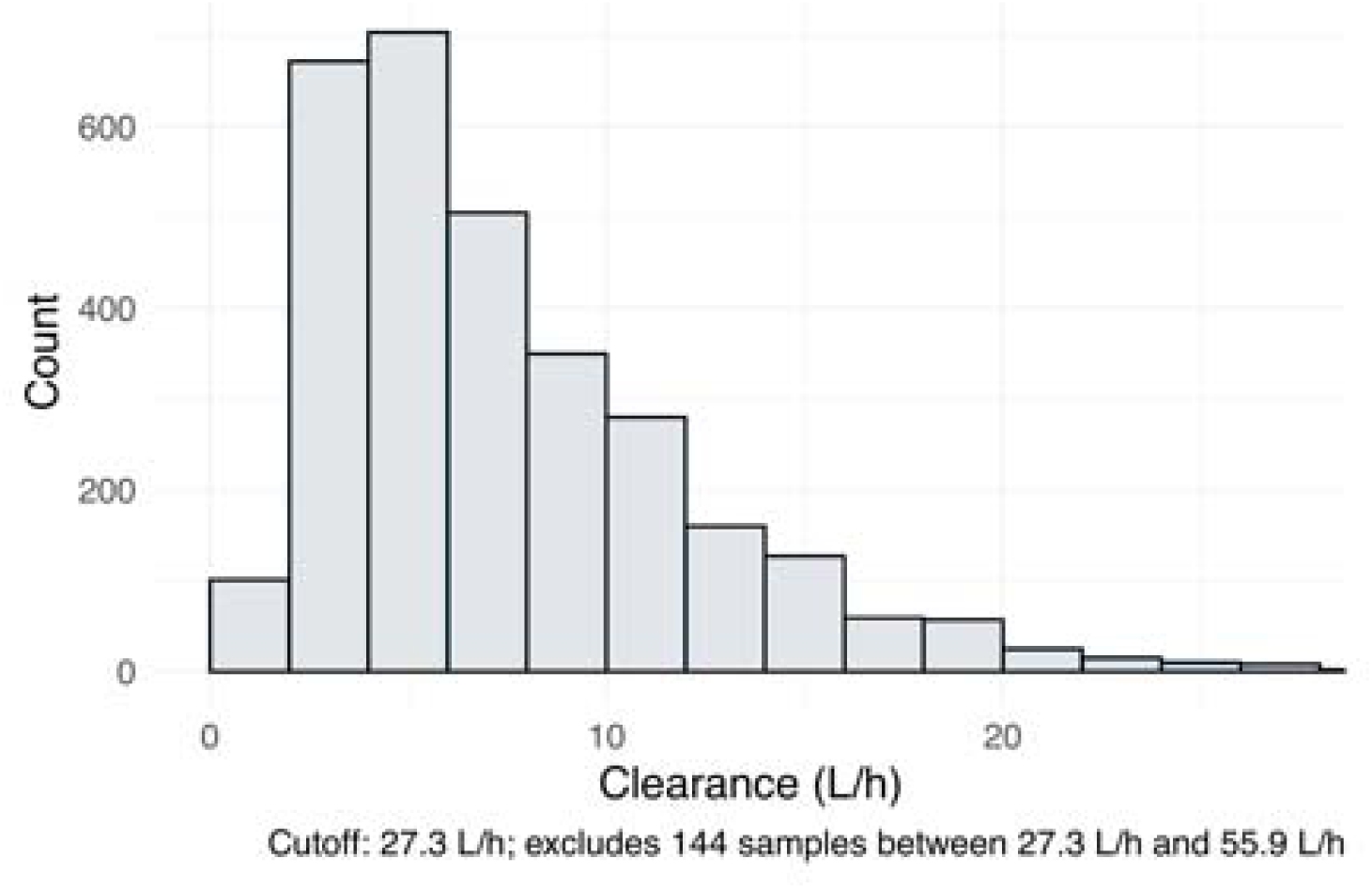
Distribution of piperacillin clearance (2 L/h bins). Values above the upper cutoff (75th percentile + 3 times IQR; 27.3 L/h) were excluded (n = 144; 4.7%). Maximum observed value: 55.9 L/h.

**Figure 2:**
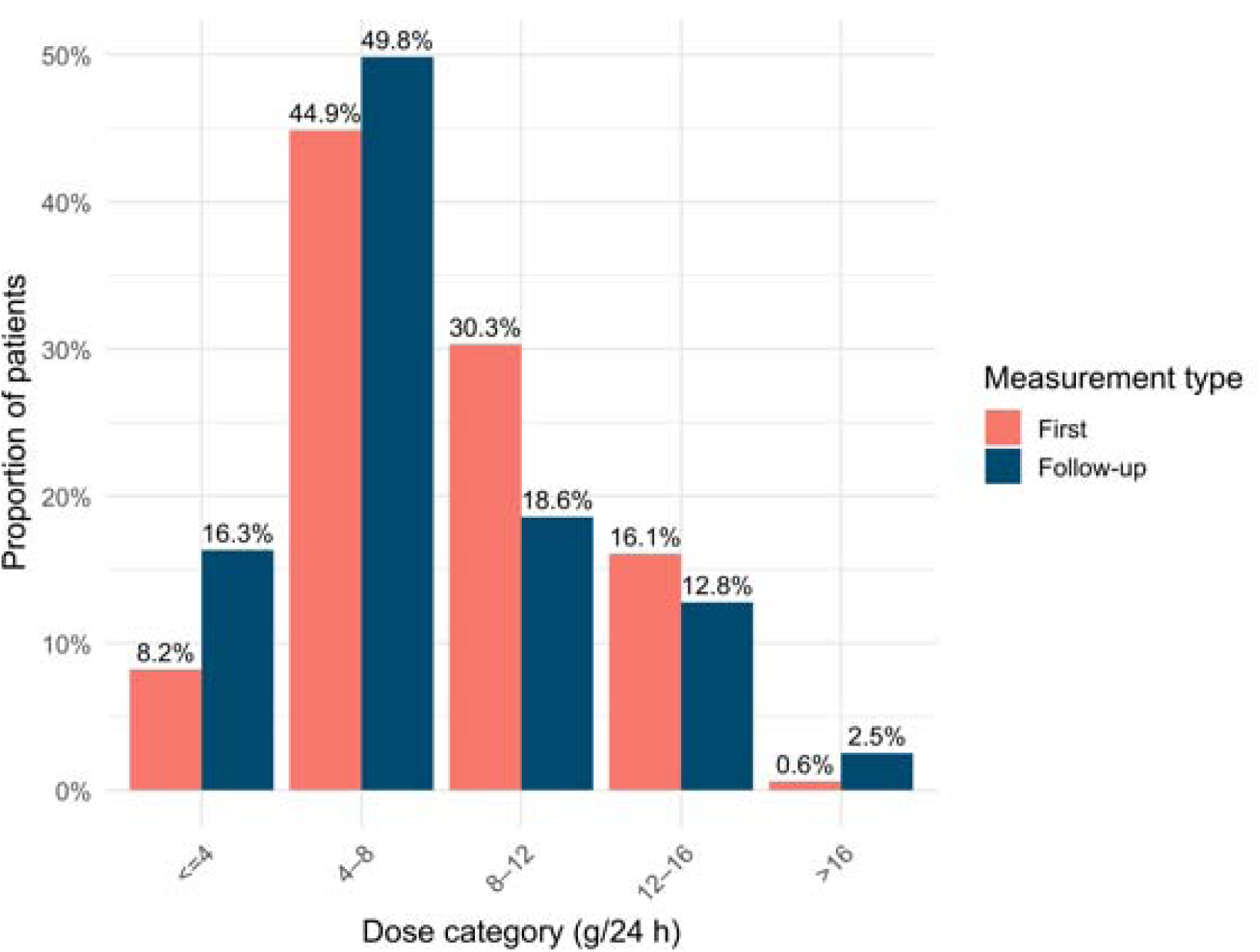
Distribution of piperacillin maintenance dose at first and follow-up (TDM-adjusted) measurements.

**Table 2:** Target attainment and distribution of piperacillin concentrations by dosing strategy.

| Dosing strategy | <32 mg/L | 32-64 mg/L<br>(therapeutic range) | 64-96 mg/L | >96 mg/L |
| --- | --- | --- | --- | --- |
| Multimodal approach<br>(First/Follow-up measurement) | 12.8%<br>(11.5%/14.1%) | 54.1%<br>(45.7%/62.4%) | 22.1%<br>(26.2%/18.0%) | 11.0%<br>(16.5%/5.6%) |
| SmPC (predicted) | 2.3% | 22.4% | 26.0% | 49.3% |
| Fixed dose 16 g<br>(predicted) | 2.2% | 20.5% | 21.1% | 56.3% |

The number of dose adjustments per patient was as follows: 0 in 951 patients, 1 in 466 patients, 2 in 100 patients, 3 in 16 patients, 4 in 5 patients, and 5 in a single patient. Dosing did differ significantly between initial and adjusted dosing (8.0 g (4.0 g – 12.1 g; 95% CI) vs. 8.0 g (3.6 g – 16.0 g; 95% CI); 1538 samples before vs. 1551 after adjustment; unpaired Wilcoxon test: p < 0.05), reflecting the significantly wider distribution spread in the adjusted group despite identical medians. Renal function was the main determinant of piperacillin clearance. Piperacillin clearance correlated significantly with creatinine clearance (r^2^ = 0.27, spearman test: ρ = 0.76, p < 0.05), while no relevant correlation was observed with body weight (r^2^ = 0.04, spearman test: p = 0.19, p < 0.05) (Figure 3). Clearance values at first measurement after therapy initiation were significantly lower to those measured later (median 5.56 L/h vs. 6.94 L/h, p < 0.05, unpaired Wilcoxon test, see web-only Supplementary Figure S3).

**Figure 3:**
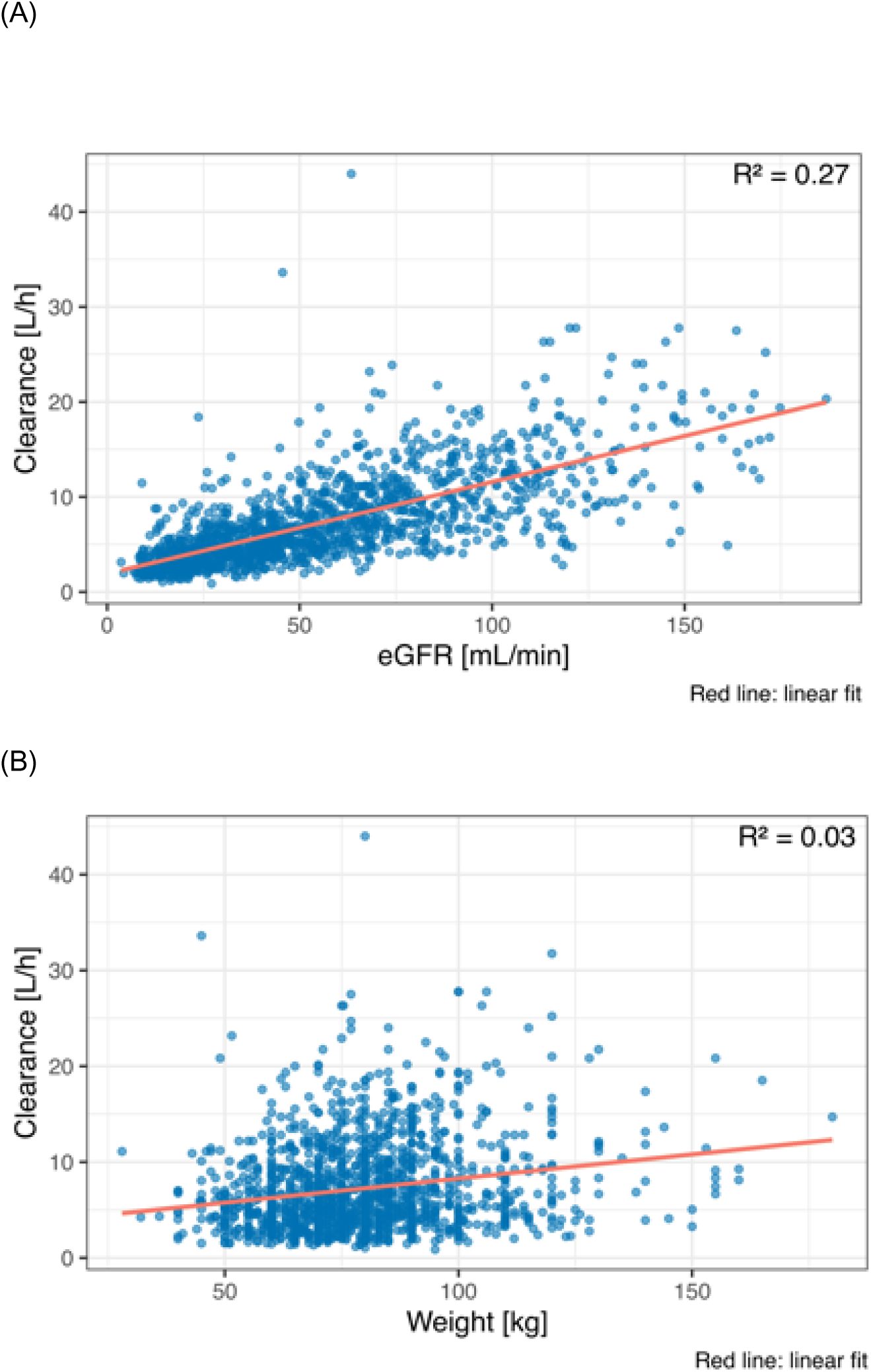
Correlation between piperacillin clearance and creatinine clearance (A) and body weight (B). Numbers indicate absolute number per group.

In addition to pharmacokinetic outcomes, individualized dosing also influenced resource utilization. Individualized continuous infusion required a median daily dose of 8 g, compared to 16 g with SmPC dosing or a fixed 16 g regimen. This corresponded to a reduction in drug consumption of 50% compared to the SmPC regimen.

### Clinical outcomes by concentration groups on initial measurement

When stratified by piperacillin exposure, ICU mortality did not appear to improve in patients with concentrations above the therapeutic range and was significantly higher in patients with moderately high concentrations (64–96 mg/L), as well as supratherapeutic concentrations (>96 mg/L). Adjustment for disease severity (SAPSII score) did not fully account for the observed association between supratherapeutic concentrations and increased mortality. Patients with subtherapeutic concentrations (<32 mg/L) showed a numerically lower, but non-significant, mortality compared with those within the therapeutic range. After adjustment for disease severity, this difference also did not suggest a clear benefit. When stratified by sex, women were approximately 1.8 times more likely to exhibit supratherapeutic piperacillin concentrations than men. However, sex itself showed only a trend and was not independently associated with mortality (OR 1.14, 95% CI 0.90–1.46). Detailed outcome data stratified by concentration group at first measurement are provided in Table 3 and illustrated in Figure 4A and 4B.

**Figure 4:**
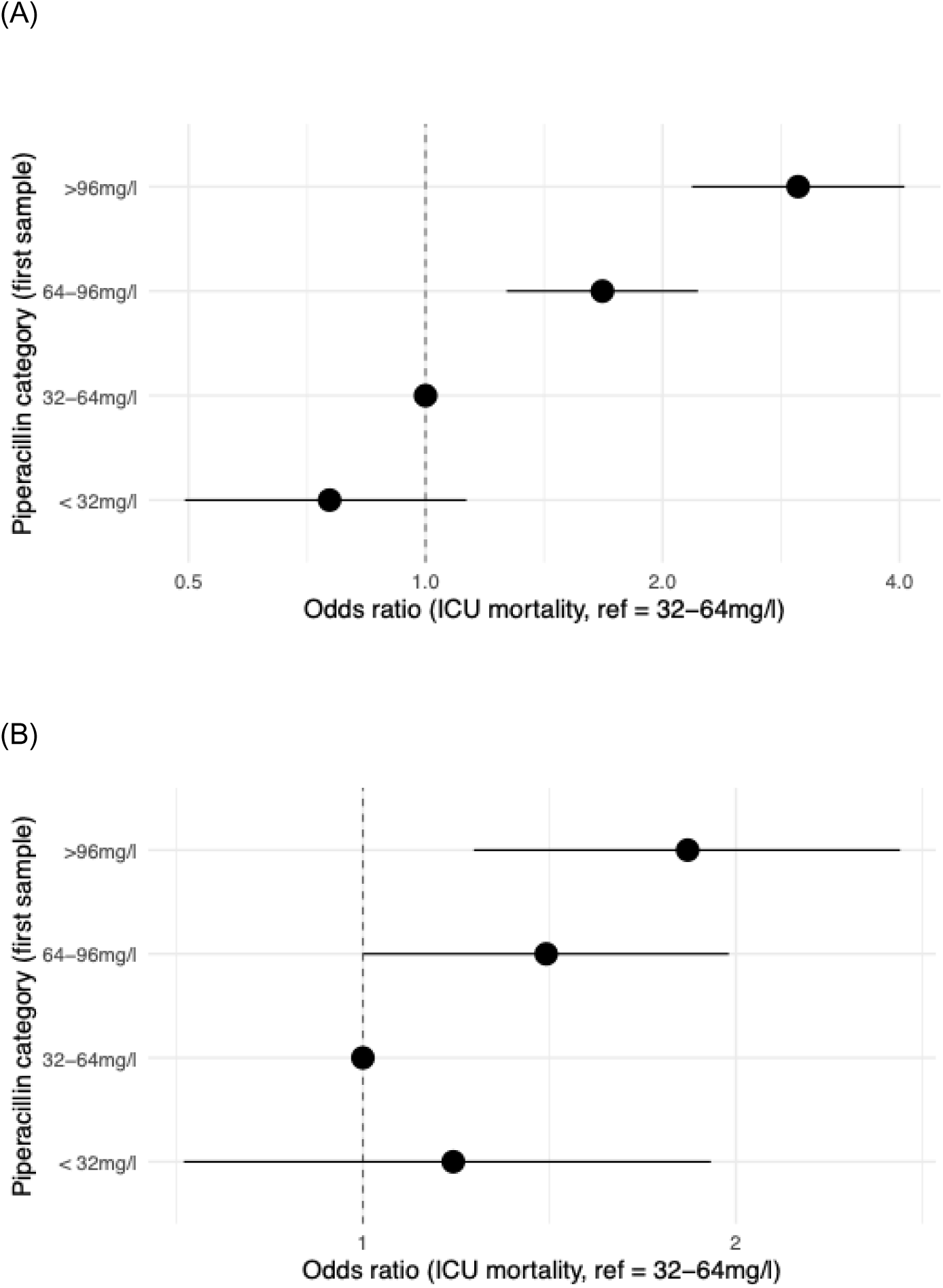
Mortality outcomes by piperacillin concentration groups (A) and adjusted for SAPS II (B). Bars represent mortality proportions; error bars indicate 95% confidence intervals.

**Table 3.** Clinical outcomes and covariates by concentration group (first concentration measurement). Numbers indicate median values; Stars show statistically significant difference between subgroup and reference group (32-64 mg/L, p < 0.05, Holmer-Bonferroni corrected).

| Concentration group | n | ICU mortality [%] | OR for ICU mortality (adj. for SAPS II) | SAPS II | TIS S-28 | BMI | Age | Piperacillin CL | eGFR (ml/min) with cutoff | Sex (male fraction, %) |
| --- | --- | --- | --- | --- | --- | --- | --- | --- | --- | --- |
| <32 mg/L | 177 | 17.2 | 1.18 (0.717, 1.91) | 32* | 10 | 28.4* | 65* | 14.0* | 77.1* | 70.7 |
| 32-64 mg/L (reference) | 704 | 21.1 | 1 (reference) | 35 | 10 | 26.7 | 72 | 8.0 | 54.7 | 69.2 |
| 64-96 | 403 | 30.6* | 1.41 (1.00, 1.97) | 39* | 10 | 25.5* | 76* | 4.4* | 39.6* | 61.6* |
| >96 mg/L | 254 | 44.5* | 1.83 (1.23, 2.71) | 40* | 10 | 25.3* | 77* | 3.3* | 33.4* | 46.5* |

BMI: body mass index; CL: clearance; eGFR: estimated glomerular filtration rate; SAPS II: Simplified Acute Physiology Score; TISS-10: Therapeutic Intervention Scoring System-10

## Discussion

This retrospective analysis of piperacillin concentrations in more than 1,500 critically ill patients demonstrates substantial PK variability. Applying a multimodal dosing approach (individualized empiric dosing, TDM-guided adjustment, and continuous infusion) led to higher target attainment than predicted for standard regimens. Importantly, very high piperacillin initial concentrations (>96 mg/L) showed a significant association with increased mortality, even after adjustment for disease severity.

Previous multicentre studies have reported up to 100-fold variability in ß-lactam exposure, mainly driven by renal function and distribution volume [2–4]. In line with these findings, we observed marked variability in our cohort with piperacillin clearance varying 85-fold and was significantly lower at therapy initiation than later in treatment. Renal clearance emerged as the main determinant of piperacillin exposure, consistent with previous evidence showing a correlation between estimated glomerular filtration rate and clearance of β-lactam antibiotics [2, 21, 22]. Based on these results, weight-based dosing or higher empiric doses during the first day of therapy should not be routinely recommended.

In addition to renal function, sex may contribute to pharmacokinetic variability through differences in body composition, protein binding, hepatic metabolism, and renal clearance. Prior work on piperacillin dose individualization indicated that women are predicted less accurately than men in published population pharmacokinetic models [23]. In our cohort, women were more likely to exhibit supratherapeutic concentrations. These aspects may warrant further investigation to clarify whether sex-related variability should be considered in dosing and dose adjustment strategies, since women are underrepresented in most pharmacokinetic studies to date.

Continuous infusion was a cornerstone of our multimodal dosing approach as proposed in the recently published BLING III trial and a subsequent meta-analysis [5, 6]. Our findings support the use of continuous infusion as a practical and effective means to stabilize drug exposure, with concentrations above the ECOFF in virtually all patients. Similar findings were reported in the TARGET trial [9], whereas studies with intermittent infusion achieved only 41-67% [2–4]. Moreover, continuous infusion facilitates TDM by allowing measurements at any time and simplifying dose adjustment. Dose individualization further enhanced target attainment in our study (46% first vs. 61% follow-up measurement). Compared with SmPC-based or fixed dosing, our multimodal approach also achieved a better balance between low and high concentrations. Subtherapeutic concentrations were corrected within 24 hours, while preventing unnecessarily high concentrations. Overall, target attainment with the multimodal approach exceeded predictions for standard or fixed dosing (54% vs. 22% vs. 21%). The TARGET trial also demonstrated significantly improved piperacillin exposure with TDM compared to the control group (37% vs. 15%) and showed fewer patients outside the target range [9]. However, these findings cannot be directly extrapolated to intermittent administration. The DOLPHIN trial demonstrated neither better clinical outcomes nor improved drug exposure, reflecting the challenges of successfully implementing such strategies in clinical practice [24].

When stratified by exposure, patients within the predefined therapeutic range (32–64 mg/L) showed the most favourable profile. Concentrations below 32 mg/L, particularly below 16 mg/L, are considered suboptimal. The trend toward a higher risk of mortality in these patients after adjustment for disease severity might underscore this consideration. Conversely, patients with very high concentrations (>96 mg/L) showed an increased risk of mortality, suggesting that more is not necessarily better for piperacillin, which is consistent with our previous 2013 analysis [15]. In TARGET, concentrations above 96 mg/L during continuous infusion were associated to a significantly higher 28-day mortality (OR 4.21, 95% CI 1.4–12.5, p = 0.01) [9]. A similar association was observed in a cohort receiving intermittent infusion when concentrations exceeded *f*T>4×MIC (OR 2.70, 95% CI 1.36–5.48, p = 0.05) [22]. Potential explanations include drug-induced toxicity, as previous reports have linked beta-lactams with neurological deterioration andpiperacillin in particular with nephrotoxicity [25–27], especially at higher exposures [26, 27]. Of note, the association persisted after SAPS II adjustment. Alternatively, high concentrations may have simply reflected underlying organ dysfunction and disease severity rather than being the direct cause of poor outcomes. Although the mechanisms remain unclear, acute kidney injury requiring renal replacement therapy is well known to be associated with poor outcomes in critically ill patients [28]. In our study, the incidence of renal replacement therapy was lower than previously reported (4% vs. 5-6%[29], with higher rates of up to 14% in patients with severe sepsis[30]), which may reflect the low number of patients with very high concentrations under the multimodal approach.

Given the retrospective design and the limited number of patients with very high concentrations, these findings should be interpreted with caution. Outcome analyses were exploratory, and residual confounding cannot be excluded despite severity adjustment. Moreover, renal clearance was estimated using the Cockcroft–Gault equation, which may not fully capture dynamic changes in critically ill patients, and non-renal clearance pathways were not assessed. However, strengths of this study include the large real-world dataset of more than 3,000 piperacillin samples collected over eight years and the multimodal dosing approach applied in clinical routine.

In clinical practice, our results emphasize the value of individualized dosing to optimize piperacillin exposure while avoiding both under- and overexposure. This aligns with international guidelines, which already recommend PK/PD-based dosing in critically ill patients [8, 31]. Broader implementation of such strategies within antimicrobial stewardship programs may contribute to more rational use of beta-lactams and potentially improve patient safety. Future research should focus on prospective multicentre trials to clarify the causal role of high concentrations. Particular attention should be given to subgroups such as patients with acute kidney injury or on renal replacement therapy in order to determine whether optimized exposure can improve hard clinical outcomes.

In conclusion, this study provides real-world evidence that a multimodal approach to piperacillin/tazobactam dosing, combining individualized empiric dosing, TDM, and continuous infusion, can ensure target attainment and help to avoid both under- and overexposure in critically ill patients. The observed indications of sex-specific differences in exposure underscore the need for further refinement of individualized dosing concepts. Beyond pharmacokinetic optimization, this approach also reduced drug use and simplified clinical workflows, potentially lowering costs and supporting its use for routine implementation.

## Author contributions

UC, OF, MW, and AB conceptualised the study. MM and SW designed the analytical workflow, and performed the statistical and pharmacokinetic analyses. Data were collected by AF, DF, MG, AR, OF, and AB. UC, MM, AR, OF, SW, AB contributed to the interpretation of data. MM was responsible for data visualisation. UC and MM drafted the manuscript. AR, OF, MW, SW, and AB critically revised the paper for important intellectual content. All authors approved the final version of the manuscript.

## Transparency declaration

UC, MM, AR, AF, OF, DF, MG, MW, SW, and AB declare that they have no conflicts of interest.

## Data Availability

All data produced in the present study are available upon reasonable request to the authors.

## Appendix A. Supplement

## References

[1] J.A. Roberts, M.H. Abdul-Aziz, J. Lipman, J.W. Mouton, A.A. Vinks, T.W. Felton, W.W. Hope, A. Farkas, M.N. Neely, J.J. Schentag, G. Drusano, O.R. Frey, U. Theuretzbacher, J.L. Kuti, P. International Society of Anti-Infective, P. the, M. Pharmacodynamics Study Group of the European Society of Clinical, D. Infectious, Individualised antibiotic dosing for patients who are critically ill: challenges and potential solutions, Lancet Infect Dis 14(6) (2014) 498–509.

[2] J.A. Roberts, S.K. Paul, M. Akova, M. Bassetti, J.J. De Waele, G. Dimopoulos, K.M. Kaukonen, D. Koulenti, C. Martin, P. Montravers, J. Rello, A. Rhodes, T. Starr, S.C. Wallis, J. Lipman, D. Study, DALI: defining antibiotic levels in intensive care unit patients: are current beta-lactam antibiotic doses sufficient for critically ill patients?, Clin Infect Dis 58(8) (2014) 1072–83.

[3] A. Abdulla, A. Dijkstra, N.G.M. Hunfeld, H. Endeman, S. Bahmany, T.M.J. Ewoldt, A.E. Muller, T. van Gelder, D. Gommers, B.C.P. Koch, Failure of target attainment of beta-lactam antibiotics in critically ill patients and associated risk factors: a two-center prospective study (EXPAT), Crit Care 24(1) (2020) 558.

[4] A.K. Smekal, M. Furebring, E. Eliasson, M. Lipcsey, Low attainment to PK/PD-targets for beta-lactams in a multi-center study on the first 72 h of treatment in ICU patients, Sci Rep 12(1) (2022) 21891.

[5] J.M. Dulhunty, S.J. Brett, J.J. De Waele, D. Rajbhandari, L. Billot, M.O. Cotta, J.S. Davis, S. Finfer, N.E. Hammond, S. Knowles, X. Liu, S. McGuinness, J. Mysore, D.L. Paterson, S. Peake, A. Rhodes, J.A. Roberts, C. Roger, C. Shirwadkar, T. Starr, C. Taylor, J.A. Myburgh, J. Lipman, B.I.S. Investigators, Continuous vs Intermittent beta-Lactam Antibiotic Infusions in Critically Ill Patients With Sepsis: The BLING III Randomized Clinical Trial, JAMA 332(8) (2024) 629–637.

[6] M.H. Abdul-Aziz, N.E. Hammond, S.J. Brett, M.O. Cotta, J.J. De Waele, A. Devaux, G.L. Di Tanna, J.M. Dulhunty, H. Elkady, L. Eriksson, M.S. Hasan, A.B. Khan, J. Lipman, X. Liu, G. Monti, J. Myburgh, E. Novy, S. Omar, D. Rajbhandari, C. Roger, F. Sjovall, I. Zaghi, A. Zangrillo, A. Delaney, J.A. Roberts, Prolonged vs Intermittent Infusions of beta-Lactam Antibiotics in Adults With Sepsis or Septic Shock: A Systematic Review and Meta-Analysis, JAMA 332(8) (2024) 638–648.

[7] F.M. Brunkhorst, M. Adamzik, H. Axer, M. Bauer, C. Bode, H.-G. Bone, T. Brenner, M. Bucher, S. David, M. Dietrich, S3-Leitlinie Sepsis–Prävention, Diagnose, Therapie und Nachsorge–Update 2025, Medizinische Klinik-Intensivmedizin und Notfallmedizin (2025) 1–69.

[8] L. Evans, A. Rhodes, W. Alhazzani, M. Antonelli, C.M. Coopersmith, C. French, F.R. Machado, L. McIntyre, M. Ostermann, H.C. Prescott, C. Schorr, S. Simpson, W.J. Wiersinga, F. Alshamsi, D.C. Angus, Y. Arabi, L. Azevedo, R. Beale, G. Beilman, E. Belley-Cote, L. Burry, M. Cecconi, J. Centofanti, A. Coz Yataco, J. De Waele, R.P. Dellinger, K. Doi, B. Du, E. Estenssoro, R. Ferrer, C. Gomersall, C. Hodgson, M. Hylander Moller, T. Iwashyna, S. Jacob, R. Kleinpell, M. Klompas, Y. Koh, A. Kumar, A. Kwizera, S. Lobo, H. Masur, S. McGloughlin, S. Mehta, Y. Mehta, M. Mer, M. Nunnally, S. Oczkowski, T. Osborn, E. Papathanassoglou, A. Perner, M. Puskarich, J. Roberts, W. Schweickert, M. Seckel, J. Sevransky, C.L. Sprung, T. Welte, J. Zimmerman, M. Levy, Surviving Sepsis Campaign: International Guidelines for Management of Sepsis and Septic Shock 2021, Crit Care Med 49(11) (2021) e1063–e1143.

[9] S. Hagel, F. Bach, T. Brenner, H. Bracht, A. Brinkmann, T. Annecke, A. Hohn, M. Weigand, G. Michels, S. Kluge, A. Nierhaus, D. Jarczak, C. Konig, D. Weismann, O. Frey, D. Witzke, C. Muller, M. Bauer, M. Kiehntopf, S. Neugebauer, T. Lehmann, J.A. Roberts, M.W. Pletz, T.T. Investigators, Effect of therapeutic drug monitoring-based dose optimization of piperacillin/tazobactam on sepsis-related organ dysfunction in patients with sepsis: a randomized controlled trial, Intensive Care Med 48(3) (2022) 311–321.

[10] H.C. Ates, A. Alshanawani, S. Hagel, M.O. Cotta, J.A. Roberts, C. Dincer, C. Ates, Unraveling the impact of therapeutic drug monitoring via machine learning for patients with sepsis, Cell Rep Med 5(8) (2024) 101681.

[11] S.G. Wicha, A.G. Martson, E.I. Nielsen, B.C.P. Koch, L.E. Friberg, J.W. Alffenaar, I.K. Minichmayr, t.P.K.P.D.s.g.o.t.E.S.o.C.M.I.D. International Society of Anti-Infective Pharmacology, From Therapeutic Drug Monitoring to Model-Informed Precision Dosing for Antibiotics, Clin Pharmacol Ther 109(4) (2021) 928–941.

[12] M. Sanz-Codina, H.O. Bozkir, A. Jorda, M. Zeitlinger, Individualized antimicrobial dose optimization: a systematic review and meta-analysis of randomized controlled trials, Clin Microbiol Infect 29(7) (2023) 845–857.

[13] S. Grau, S. Luque, O. Ferrandez, A. Benitez Cano, D. Rubio-Rodriguez, C. Rubio-Terres, Economic impact of individualized antimicrobial dose optimization in the critically ill patient in Spain, Front Pharmacol 16 (2025) 1506109.

[14] U. Chiriac, O.R. Frey, A.C. Roehr, A. Koeberer, P. Gronau, T. Fuchs, J.A. Roberts, A. Brinkmann, Personalized ss-lactam dosing in patients with coronavirus disease 2019 (COVID-19) and pneumonia: A retrospective analysis on pharmacokinetics and pharmacokinetic target attainment, Medicine (Baltimore) 100(22) (2021) e26253.

[15] U. Chiriac, D.C. Richter, O.R. Frey, A.C. Rohr, S. Helbig, J. Preisenberger, S. Hagel, J.A. Roberts, M.A. Weigand, A. Brinkmann, Personalized Piperacillin Dosing for the Critically Ill: A Retrospective Analysis of Clinical Experience with Dosing Software and Therapeutic Drug Monitoring to Optimize Antimicrobial Dosing, Antibiotics 10(6) (2021) 667.

[16] EUCAST. Breakpoints Piperacillin. Available online: http://www.eucast.org/clinical_breakpoints (accessed on 09 December 2025).

[17] D. Czock, V. Schwenger, D. Kindgen-Milles, M. Joannidis, S. John, M. Schmitz, A. Jorres, A. Zarbock, M. Oppert, J.T. Kielstein, C. Willam, [Dose adjustment of anti-infective drugs in patients with renal failure and renal replacement therapy in intensive care medicine : Recommendations from the renal section of the DGIIN, OGIAIN and DIVI], Med Klin Intensivmed Notfmed 113(5) (2018) 384–392.

[18] Fresenius Kabi. Summary of Product Characteristics. Piperacillin/Tazobactam Kabi 4 g/0,5 g. Available online: https://www.fachinfo.de/fi/pdf/011759 (accessed on 09 December 2025).

[19] R.P. Dellinger, M.M. Levy, A. Rhodes, D. Annane, H. Gerlach, S.M. Opal, J.E. Sevransky, C.L. Sprung, I.S. Douglas, R. Jaeschke, T.M. Osborn, M.E. Nunnally, S.R. Townsend, K. Reinhart, R.M. Kleinpell, D.C. Angus, C.S. Deutschman, F.R. Machado, G.D. Rubenfeld, S. Webb, R.J. Beale, J.L. Vincent, R. Moreno, S. Surviving Sepsis Campaign Guidelines Committee including The Pediatric, Surviving Sepsis Campaign: international guidelines for management of severe sepsis and septic shock, 2012, Intensive Care Med 39(2) (2013) 165–228.

[20] A.C. Roehr, O.R. Frey, A. Koeberer, T. Fuchs, J.A. Roberts, A. Brinkmann, Anti-infective drugs during continuous hemodialysis - using the bench to learn what to do at the bedside, Int J Artif Organs 38(1) (2015) 17–22.

[21] J.J. De Waele, J. Lipman, M. Akova, M. Bassetti, G. Dimopoulos, M. Kaukonen, D. Koulenti, C. Martin, P. Montravers, J. Rello, A. Rhodes, A.A. Udy, T. Starr, S.C. Wallis, J.A. Roberts, Risk factors for target non-attainment during empirical treatment with beta-lactam antibiotics in critically ill patients, Intensive Care Med 40(9) (2014) 1340–51.

[22] S. Drager, T.M.J. Ewoldt, A. Abdulla, W.J.R. Rietdijk, N.J. Verkaik, P. van Vliet, I.M. Purmer, M. Osthoff, B.C.P. Koch, H. Endeman, D. investigators, Target attainment of beta-lactam antibiotics and ciprofloxacin in critically ill patients and its association with 28-day mortality, J Crit Care 85 (2025) 154904.

[23] S. Greppmair, A. Brinkmann, A. Roehr, O. Frey, S. Hagel, C. Dorn, A. Marsot, I. El-Haffaf, M. Zoller, T. Saller, J. Zander, L.M. Schatz, C. Scharf, J. Briegel, I.K. Minichmayr, S.G. Wicha, U. Liebchen, Towards model-informed precision dosing of piperacillin: multicenter systematic external evaluation of pharmacokinetic models in critically ill adults with a focus on Bayesian forecasting, Intensive Care Med 49(8) (2023) 966–976.

[24] T.M.J. Ewoldt, A. Abdulla, P. van den Broek, N. Hunfeld, S. Bahmany, A.E. Muller, D. Gommers, S. Polinder, H. Endeman, I. Spronk, B.C.P. Koch, Barriers and facilitators for therapeutic drug monitoring of beta-lactams and ciprofloxacin in the ICU: a nationwide cross-sectional study, BMC Infect Dis 22(1) (2022) 611.

[25] B. Navalkele, J.M. Pogue, S. Karino, B. Nishan, M. Salim, S. Solanki, A. Pervaiz, N. Tashtoush, H. Shaikh, S. Koppula, J. Koons, T. Hussain, W. Perry, R. Evans, E.T. Martin, R.P. Mynatt, K.P. Murray, M.J. Rybak, K.S. Kaye, Risk of Acute Kidney Injury in Patients on Concomitant Vancomycin and Piperacillin-Tazobactam Compared to Those on Vancomycin and Cefepime, Clin Infect Dis 64(2) (2017) 116–123.

[26] S. Imani, H. Buscher, D. Marriott, S. Gentili, I. Sandaradura, Too much of a good thing: a retrospective study of beta-lactam concentration-toxicity relationships, J Antimicrob Chemother 72(10) (2017) 2891–2897.

[27] Y. Zerbib, C. Gaulin, S. Bodeau, B. Batteux, A.S. Lemaire-Hurtel, J. Maizel, L. Kontar, Y. Bennis, Neurological burden and outcomes of excessive beta-lactam serum concentrations of critically ill septic patients: a prospective cohort study, J Antimicrob Chemother 78(11) (2023) 2691–2695.

[28] S.M. Bagshaw, S. Uchino, R. Bellomo, H. Morimatsu, S. Morgera, M. Schetz, I. Tan, C. Bouman, E. Macedo, N. Gibney, A. Tolwani, H.M. Oudemans-van Straaten, C. Ronco, J.A. Kellum, Beginning, I. Ending Supportive Therapy for the Kidney, Septic acute kidney injury in critically ill patients: clinical characteristics and outcomes, Clin J Am Soc Nephrol 2(3) (2007) 431–9.

[29] V.N.A.R.F.T. Network, P.M. Palevsky, J.H. Zhang, T.Z. O’Connor, G.M. Chertow, S.T. Crowley, D. Choudhury, K. Finkel, J.A. Kellum, E. Paganini, R.M. Schein, M.W. Smith, K.M. Swanson, B.T. Thompson, A. Vijayan, S. Watnick, R.A. Star, P. Peduzzi, Intensity of renal support in critically ill patients with acute kidney injury, N Engl J Med 359(1) (2008) 7–20.

[30] A. Sakhuja, G. Kumar, S. Gupta, T. Mittal, A. Taneja, R.S. Nanchal, Acute Kidney Injury Requiring Dialysis in Severe Sepsis, Am J Respir Crit Care Med 192(8) (2015) 951–7.

[31] M.H. Abdul-Aziz, J.C. Alffenaar, M. Bassetti, H. Bracht, G. Dimopoulos, D. Marriott, M.N. Neely, J.A. Paiva, F. Pea, F. Sjovall, J.F. Timsit, A.A. Udy, S.G. Wicha, M. Zeitlinger, J.J. De Waele, J.A. Roberts, M. Infection Section of European Society of Intensive Care, Pharmacokinetic/pharmacodynamic, M. Critically Ill Patient Study Groups of European Society of Clinical, D. Infectious, M. Infectious Diseases Group of International Association of Therapeutic Drug, T. Clinical, I.C.U. Infections in the, C. Sepsis Working Group of International Society of Antimicrobial, Antimicrobial therapeutic drug monitoring in critically ill adult patients: a Position Paper(), Intensive Care Med 46(6) (2020) 1127–1153.

