## Supplementary material for "Pharmacokinetics, target attainment and outcomes of piperacillin/tazobactam in critically ill patients receiving continuous infusion with therapeutic drug monitoring: a retrospective analysis": Online supplement

### Methods

Concentrations were predicted using the observed piperacillin clearance ($CL_{PIP}$). $CL_{PIP}$ was calculated from the observed piperacillin concentrations in steady-state $cp_{PIP}$ and the utilised infusion rate ($R_{inf}$) (Equation 1).

Equation 1:

$$CL_{PIP} = \frac{R_{inf}}{cp_{PIP}}$$

For the target attainment analysis for SmpC and fixed dose regimens (Table 2), piperacillin concentrations were calculated accordingly (Equation 2).

Equation 2:

$$cp_{PIP} = \frac{R_{inf}}{CL_{PIP}}$$

For calculation of eGFR from creatinine concentrations, the Cockcroft-Gault equation was used (Equation 3):

$$eGFR = \frac{(140 - Age)}{Serum\ creatinine} * \frac{Weight}{72} * 0.85\ (if\ female)$$

Serum creatinine was limited to 0.6 for women and 0.7 for men (lower bound). When the actual patient weight (BW) exceeded the ideal body weight (IBW), the ideal body weight +40% of the difference between ideal and actual bodyweight was used (Equation 4):

$$Weight = 0.4 * (WT_{real} - WT_{ideal}) + WT_{ideal}\ ; if\ BW > IBW$$

Ideal bodyweight was calculated according to the Devine equation (Equation 5):

$$IBW_{male} = 50kg + 0.91kg/cm * (Height[cm] - 152.4)$$

$$IBW_{female} = 45.5kg + 0.91kg/cm * (Height[cm] - 152.4)$$

**Figure S1:** Distribution of piperacillin concentrations under the multimodal approach.

Vertical lines indicate cutoffs (16 mg/L, 32 mg/L, 64 mg/L, 96 mg/L).

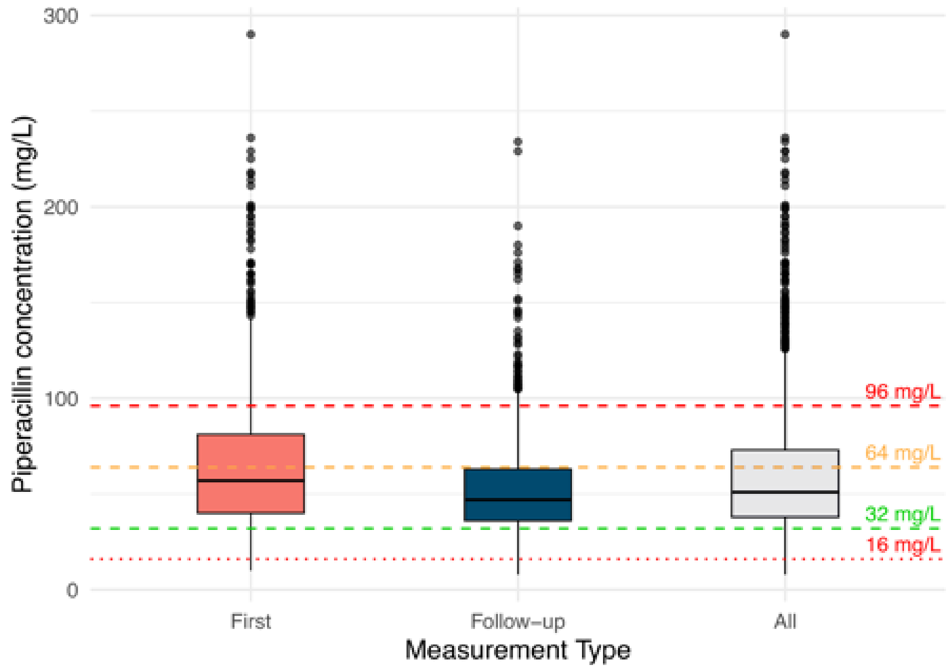


**Figure S2:** Target attainment and distribution of piperacillin concentrations under the multimodal approach (first, follow-up, all measurements). Numbers on bars indicate absolute count.

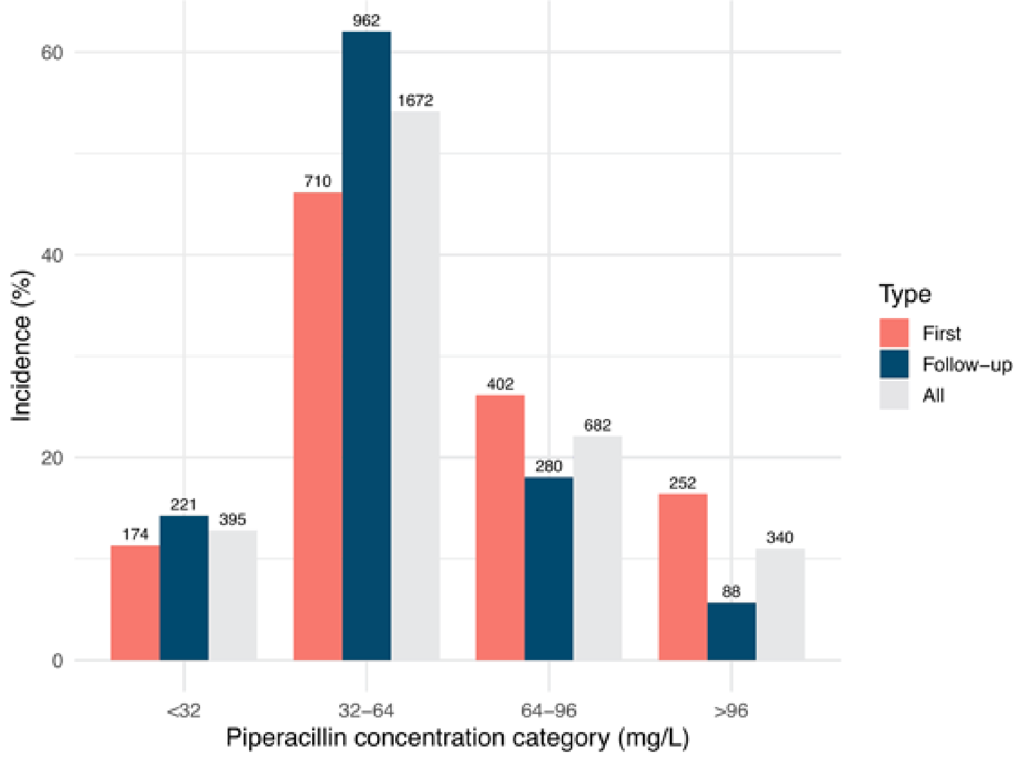

**Figure S3:** Piperacillin clearance by sample type.

Piperacillin clearance stratified by first measurement vs. follow-up measurement after dose adjustment vs. all measurements.

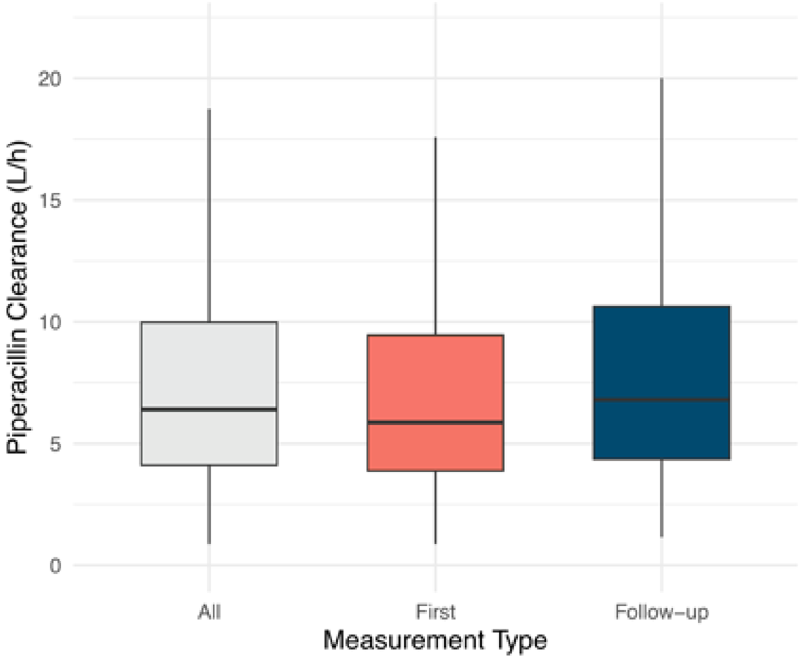
